# Systematic review of randomised controlled clinical trials examining effectiveness of contingent financial rewards for smoking cessation during pregnancy: intention-to-treat and causal effects on birthweight

**DOI:** 10.1101/2024.08.21.24312341

**Authors:** David Tappin, Jiyoung Lee, Alex McConnachie, Loren Kock, Stephen T. Higgins, Sarah H. Heil, Ivan Berlin, Steven J. Ondersma, Frank Kee, Ira Bernstein, John Van Sicklen Maeck, Linda Bauld

## Abstract

**BACKGROUND:** *Objective:* To examine birth weight change caused by adding financial rewards for smoking cessation compared to no rewards for pregnant women. To estimate the average expected birth weight change for those who quit because of rewards.

**METHODS:** This study updates a previous systematic review and refocuses the outcome from smoking cessation to birth weight.

*Eligibility Criteria:* Trials with an experimental design allowing treatment effects to be attributed to rewards were included. Trials involving non-pregnant participants, or with no report of magnitude, treatment duration, timing or where most rewards were contingent on another behaviour (e.g., treatment attendance) were excluded.

*Information sources:* Medline, PsycInfo, Embase, Cochrane (Central Register of Controlled Trials, Tobacco Addiction Group Specialised Register and Database of Systematic Reviews), and PubMed searched to 5th December 2023.

*Risk of bias:* Risk of bias and certainty of evidence used Cochrane ‘Risk of bias 2’ and GRADE assessments.

*Synthesis of results:* Primary analysis estimated Intention-To-Treat (ITT) mean birthweight difference when randomised to offer of rewards versus control. Within-trial estimates and standard errors were derived from mean, standard deviation, and sample size data provided, or from publications. Pooled ITT estimates used common (fixed) and random effects models. Secondary analyses used trial team supplied data to derive Complier Average Causal Effect (CACE) estimate of smoking cessation on birth weight, and a standard error. Estimates were pooled using common and random effects models. Similar analyses were applied to low birth weight (<2500g), birth weight for gestational age z-scores, and small for gestational age (<10^th^ percentile).

**RESULTS:** *Included studies:* Primary analysis included 8 trials (2351 participants) from the UK (2 trials, 1475 participants); France (1 trial, 407 participants), and the US (6 trials, 469 participants). Secondary analysis included 7 trials as data retrieval from one US trial (51 participants) was not possible.

*Synthesis of results:* Primary ITT analysis (2351 participants) estimated a mean 46.3g (95% CI: 0.0 to 92.6) birth weight increase when offered financial rewards for smoking cessation. Secondary CACE analysis (2239 participants) estimated a mean 206.0g (95% CI: -69.1 to 481.1) increase for smokers who quit because of rewards. There was no effect on low birth weight (<2500g), or birth weight adjusted for gestational age, though less babies were born small for gestational age, particularly if cessation was because of rewards (CACE risk difference -17.7%; 95% CI: -34.9% to -0.4%).

**DISCUSSION:** *Limitation of evidence:* Sample size led to *imprecision* - maximum 2351 participants. A single trial of 3712 participants would give 80% power at 5% significance to show a 46g increase from 3.1kg to 3.146kg with 0.5kg standard deviation in both groups. *Consistency* - trials where smoking cessation increased (7 of 8) all showed a mean birth weight increase. In one trial smoking cessation fell as did birth weight. *Bias* is unlikely as 3 of 4 trials with no birth weight data showed increased cessation *consistent* with higher mean birth weight.

*Interpretation:* Trials of contingent financial rewards for smoking cessation have previously been shown to more than double pregnancy quit rates. We have uncovered a significant (46g) population level increase in mean birth weight, driven by a clinically important mean increase (206g) for those who quit because of financial rewards associated with a reduction in Small for Gestational Age births.

**OTHER:** *Funding:* Review update - The U.S. National Institute of Health, National Institute of General Medical Sciences Center of Biomedical Research Excellence Award P30GM149331. Data retrieval, synthesis and analysis – Scottish Cot Death Trust.

*Registration:* https://www.crd.york.ac.uk/prospero/display_record.php?ID=CRD42024494262

## Rationale

Smoking throughout pregnancy is one of the most damaging behaviours affecting the unborn child associated with a 10% decrease in birth weight (mean 387g) ^1^ for ‘consistent smoking’ and many other short- and long-term problems causing families great distress ^2, 3^ as well as large additional health service costs ^4, 5^. A 10% decrease in birth weight particularly if associated with being Small for Gestational Age (SGA) is a physical marker of significant and often long-term damage ^6^. Avoidance of this birth weight reduction caused by smoking during pregnancy would be a worthwhile and clinically effective intervention that could help to convince policy makers to implement financial rewards to help pregnant women who smoke to quit throughout the UK and beyond.

### Systematic review and meta-analysis of smoking cessation towards the end of pregnancy

The addition of financial rewards has been shown to be the most effective intervention to increase the effectiveness of Stop Smoking Services (SSS) for pregnant women ^7, 8^. In this paper SSS is used as a generic term for any support given to help women to stop smoking during pregnancy. Generally, only those compliant with quitting smoking during pregnancy receive financial rewards. More than half of the trials^8^ also reported birthweight with most showing a non-significant increase in birth weight for those offered financial rewards for smoking cessation compared to no financial reward. This study aims to assess if the addition of financial rewards to usual care significantly increases infant birth weight compared to usual care alone.

### Complier Average Causal Effects (CACE) meta-analysis of birth weight improvement

The effect of ‘consistent smoking’ – an average 387g reduction in birthweight ^1^ is much greater than the mean improvement in birthweight found in the trials of financial rewards. An example is the CPIT II trial where the improvement was 21g ^9^. This difference largely relates to compliance. Trials of conditional rewards for smoking cessation in pregnancy are examples of an encouragement trial. All participants are free to stop smoking or not, but those randomised to the intervention arm receive additional encouragement to do so; in this case through the offer of a financial reward if they are successful. For some people, the intervention makes no difference, and they cannot stop smoking either with or without the encouragement. At the other end of the spectrum, there are those who can quit without the intervention, and though the intervention results in a reward, it actually has no effect on their ability to stop smoking. Only those in the middle, who are not able to quit without the intervention, but are able to do so with additional encouragement, are affected by the intervention. Logically, it is only this group of people who stand to achieve downstream health benefits, for example in terms of the birthweight of their child.

With such a study design, Complier Average Causal Effects (CACE) analysis can be used to estimate the effect of stopping smoking on birth weight ^10-12^. In a randomized encouragement trial, CACE analysis estimates the effect of the behaviour change (stopping smoking) on the outcome (birthweight) in those people who achieve the behaviour change (smoking cessation) only as a result of the randomised encouragement intervention (the offer of financial rewards).

In the trial Tappin et al 2015 ^9^, the proportion of pregnant participants who quit smoking towards the end of pregnancy was 8.6% in the usual Stop Smoking Service control group which increased to 22.5% with the additional offer of financial rewards for smoking cessation. CACE analysis ^10-12^ indicated that the ‘small’ birth weight improvement of 21g in those offered financial rewards as well usual Stop Smoking Service care compared to those offered usual care alone, translated into a 154g (95% CI: -617 to 803) (about 5% of birthweight) improvement for those women who quit smoking but would not have managed to quit without the additional offer of financial rewards. However, the overall 21g increase in birthweight and this clinically important 154g increase among those affected did not reach statistical significance and have therefore largely been ignored by clinicians and policymakers.

This current paper extends the systematic review by Kock et al ^8^ and focuses the outcome on birth weight when the offer of financial rewards for smoking cessation is added to routine smoking cessation support for pregnant women. All corresponding authors for the studies included in the Kock review and update were invited to provide additional data to allow a meta-analysis of the population-level impact of the offer of financial rewards on birth weight, and of the causal effect of smoking cessation in the subset of women who were able to quit as a result of the intervention.

#### Research questions

1. Do babies born to women who smoke in early pregnancy show an increase in mean birth weight and mean birth weight for gestational age z-score if women are offered financial rewards contingent to quitting smoking as well as usual Stop Smoking Service support compared with babies born to mothers offered usual Stop Smoking Service support alone or rewards not contingent on smoking cessation (Intention to Treat meta-analysis)?
2. What is the mean birthweight difference associated with quitting smoking during pregnancy as a result of the offer of financial rewards? How is this reflected in numbers of babies born low birth weight <2500g and Small for Gestational Age <10th percentile?

## Methods

We performed a systematic review and meta-analyses of randomised controlled trials examining the impact of contingent financial rewards for smoking cessation during pregnancy on birth weight. This study is based on the systematic review by Kock et al.^8^ The study is reported according to the preferred reporting items for systematic reviews and meta-analyses (PRISMA) guidelines ^13^. The study protocol was registered on PROSPERO (international prospective register of systematic reviews) and is available online (www.crd.york.ac.uk/prospero, CRD42024494262).

### PROTOCOL

Change was made to the protocol (www.crd.york.ac.uk/prospero, CRD42024494262) before data were collected after discussion with lead authors of trials in Kock.^8^ Low Birth Weight <2.5kg and Small for Gestational Age were suggested as additional outcome variables to be examined. Changes were made to data collection and analysis methods.

### SEARCH STRATEGY AND SELECTION CRITERIA

The search strategy and selection criteria followed that of Kock 2023 ^8^ (registered PROSPERO ID CRD42022372291) but was updated to allow inclusion of studies that have been published after the cutoff of the Kock review (specifically between 17^th^ November 2022 and 5^th^ December 2023) and followed PRISMA guidelines ^13^.

### STUDY SELECTION

Study screening and selection for the Kock et al review is reported elsewhere ^8^. For the current review update, potentially relevant studies retrieved from the updated search were screened by LK, with detailed reasons for exclusion reported in Appendix 1.

### DATA EXTRACTION

We reviewed studies identified in the updated search by Kock et al ^8^. For studies that did not report birth weight, we contacted the corresponding authors to provide this information. Data were requested for birth weight and birth weight for gestational age z-score (calculated by the corresponding authors using https://intergrowth21.tghn.org/standards-tools/ ^14^), including sample size, mean and standard deviation, and the number of births (<2.5kg for birth weight data and <10th percentile z-score, <-1.2816 for birth weight for gestational age z-score data, respectively). For CACE analyses, where available, these data were also collected for four subgroups defined by randomised group and smoking status (whether participants stopped or continued smoking).

### DATA SYNTHESIS AND ANALYSIS

The intention-to-treat (ITT) effects were estimated by the differences in means (for birth weight and birth weight z-score) and risks (for low birth weight (<2500g)^15^ and small for gestational age (<10^th^ percentile)) between randomised groups. The complier average causal effects (CACE) were estimated using instrumental variable regression models. The monotonicity assumption (no defiers) for CACE analysis was evaluated by examining the estimated compliance rates. The ITT and CACE estimates were then pooled under both fixed and random effects models. The pooled effect estimates were expressed as mean differences for continuous outcomes and risk differences for dichotomous outcomes, with 95% confidence intervals. Heterogeneity was examined by estimates of between study variance (τ²) and the I² statistics. The possibility of publication bias was not assessed as fewer than ten trials were included in all analyses. Cumulative meta-analyses based on publication date were conducted to evaluate evidence accumulation. Influential studies were assessed using the leave-one-out method to assess the sensitivity of the overall results. Meta-analyses were done using ‘meta’ package in R (version 4.2.1)^16^.

### RISK OF BIAS

The GRADE system ^17^ was used to describe the strength or weakness of recommendations emanating from the findings of this systematic review.

### ETHICS APPROVAL

The PROSPERO protocol ^18^ was reviewed by the West of Scotland National Health Service research ethics manager January 3^rd^ 2024, and full application was submitted to the College of Medical, Veterinary and Life Sciences ethics committee at Glasgow University 20^th^ March 2024. Both determined that the study could go ahead.

### FUNDING

Funding for data analysis utilised residual contingency funding from the CPIT III trial ^19^ with permission from the funder – the Scottish Cot Death Trust November 3, 2023. The U.S. National Institute of General Medical Sciences Center of Biomedical Research Excellence Award P30GM149331 provided support for Dr Kock to update the systematic review from 17^th^ November 2022 to 5^th^ December 2023.

### PATIENT AND PUBLIC INVOLVEMENT

Patient and public involvement was not undertaken for this updated systematic review and meta-analysis.

## Results

Further searches by Kock to 5^th^ December 2023 found 10 additional studies to be assessed. Details of these studies and reasons all were excluded are described in Appendix 1.

Therefore, our meta-analyses are based on the trials included in the Kock review ^8^; 12 studies with a combined relative risk of smoking cessation towards the end of pregnancy of 2.43 (95% CI: 2.04 to 2.91). Details of the studies are available in Kock ^8^ table 1.

**Table 1:**
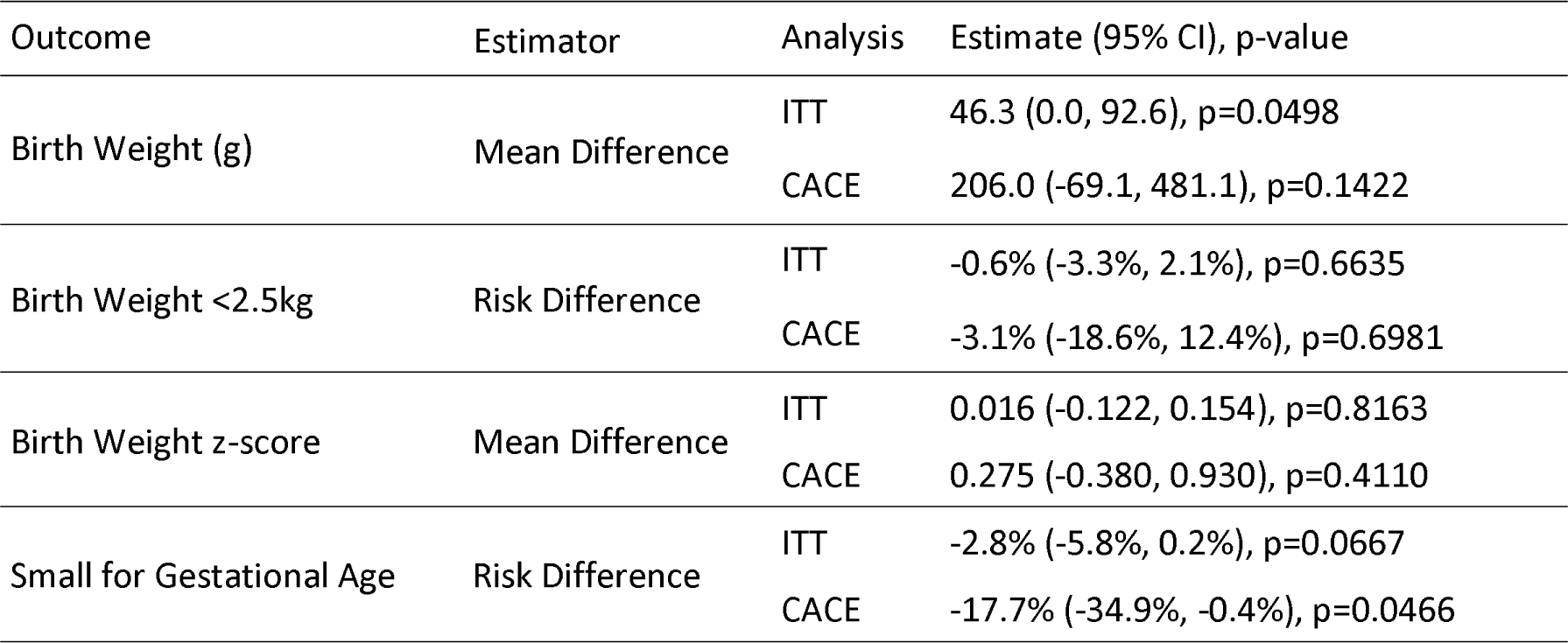
Pooled estimates of the Intention-To-Treat (ITT) effects of the offer of financial rewards, and Complier Average Causal Effect (CACE) effects of smoking cessation, on study outcomes. Pooled estimates derived from common (fixed) effects models.

Assessments of risk of bias for individual trials in the Kock review ^8^ are available at supplementary appendix: https://www.sciencedirect.com/science/article/pii/S0091743523002347

CACE analyses were subject to the availability of subgroup data and that met the required conditions for CACE analysis. One trial showed a negative estimated compliance rate, indicating the possible presence of defiers; therefore, this trial was excluded from the CACE analyses.

The common and random effects models gave very similar results, so the common effects models are reported in the figures. Table 1 gives the pooled effect estimates from all common effects models. Supplementary Table 1 shows the corresponding results from random effects models.

### Birth weight

Data on birth weight were available for eight trials,^9, 19-25^ including a total of 2351 participants. Pooled analysis showed that the average birth weight of babies born to women in the financial rewards group was 46.3g higher (95% CI: 0.0 to 92.6; GRADE=moderate) compared with the control group (Figure 1; Table 1). In the CACE analysis, we included six trials ^9, 19-23^ based on the availability of subgroup data on birth weight that met the required conditions for the analysis. Pooled CACE estimate showed that smoking cessation due to financial rewards resulted in an average gain of 206.0g in newborn weight (95% CI: -69.1 to 481.1; n=2239), which was, however, statistically non-significant (Figure 2; Table 1). A sensitivity analysis including a trial that did not meet the monotonicity condition for CACE analysis showed similar results (Supplementary Figure 11).

**Figure 1:**
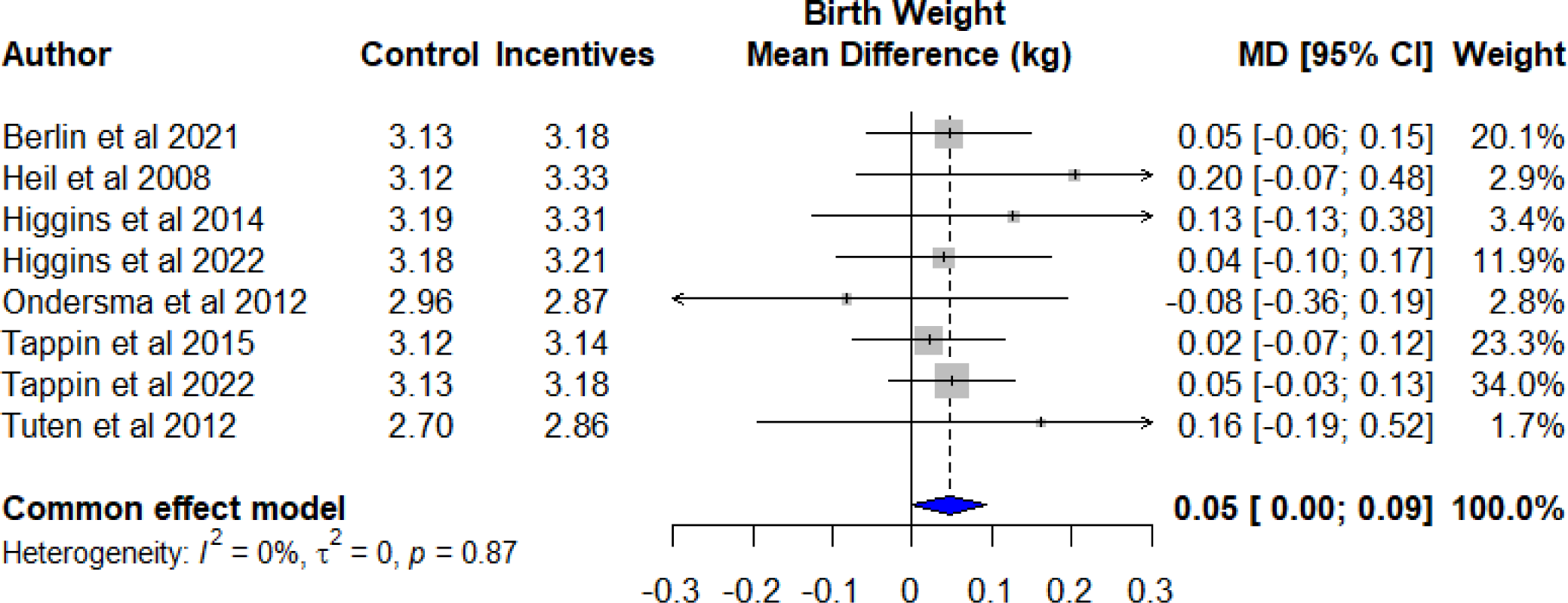
Intention-to-treat (ITT) estimates of the effect of the offer of financial rewards for smoking cessation during pregnancy on birth weight (kg). The size of data markers is proportional to the weight in the meta-analysis.

**Figure 2:**
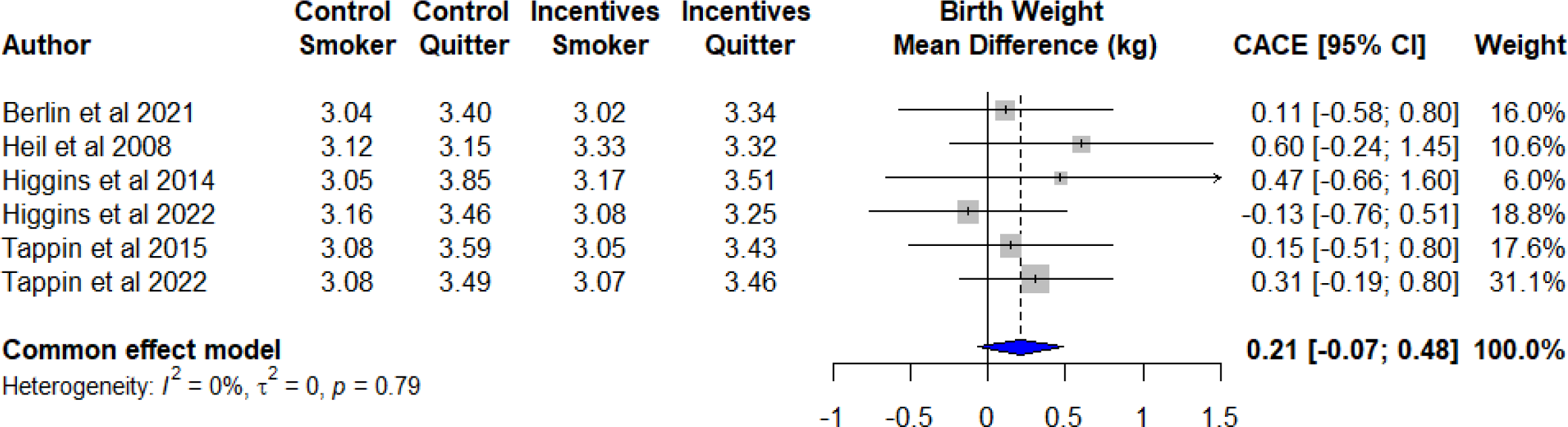
Complier Average Causal Effect (CACE) estimates of the effect of smoking cessation during pregnancy on birth weight (kg). The size of data markers is proportional to the weight in the meta-analysis.

The GRADE approach^17^ was used to systematically assess the certainty of evidence that birth weight increases with the offer of financial rewards for smoking cessation to pregnant women (appendix 2). Although statistically significant, evidence for the increase in birthweight was graded as moderate due to potential imprecision in the effect estimate (95% CI 0 to 93g improvement), likely related to sample size.

### Low birth weight (< 2.5kg)

Pooling showed that there was no clear evidence of ITT effect (risk difference -0.6%; 95 CI: - 3.3% to 2.1%; 7 studies; n=2300) or CACE (risk difference -3.1%; 95% CI: -18.6% to 12.4%; 6 studies; n=2239) on the risk of low birth weight delivery (Supplementary Figure 1-2; Table 1).

### Birth weight for gestational age z-score and small for gestational age

To account for variations in birth weight with gestational age and sex, we performed meta-analyses on birth weight z-scores. Data on birth weight z-scores were available from five trials ^20-23, 25^. Pooled effect estimates are reported in Table 1 and illustrated in Supplementary Figure 3-6. Our pooled analyses showed no clear evidence of ITT effect or CACE on birth weight adjusted for gestational age and sex.

We also investigated the effect of financial rewards on the risk of babies born small for gestational age (z-score <10^th^ percentile). Pooling showed a small and non-significant reduction in the risk of being born for Small for Gestational Age(SGA) with the offer of financial rewards alone (risk difference 2.8%; 95% CI: -5.8% to 0.2%; 5 studies; n=825) (Supplementary Figure 5; Table 1). However, we found some evidence of a significant reduction in the risk of SGA due to smoking cessation induced by the rewards, with our pooled CACE analysis showing a 17.7% reduced risk (95% CI: -34.9% to -0.4%; 4 studies; n=755) in those who quit smoking as a result of the intervention. (Supplementary Figure 6; Table 1).

### Sensitivity Analyses

Supplementary Figures 7 and 8 show cumulative forest plots for each analysis. As more data have accrued the pooled estimates appear to have stabilized, and become more precise. Supplementary figures 9 and 10 show pooled estimates for each analysis after leaving each trial out in turn. With little heterogeneity observed overall, no individual trial appears to be having an undue influence on any of the analyses.

## Discussion

This systematic review and meta-analysis found a significant increase in birth weight of 46.3g (95% CI: 0.0 to 92.6) when pregnant women who smoke were offered the addition of financial rewards contingent on smoking cessation during pregnancy compared to those offered routine smoking cessation support alone (whatever that support may be).

These data also provide a best estimate for the increase in birth weight associated with smoking cessation during pregnancy because of financial rewards of 206.0g (95% CI: -69.1 to 481.1) or 6.1% of average birth weight (3.4kg), less than the estimate of 387g reduction for smokers from a recent cohort study ^1^. One explanation for this difference is that our estimate is from randomised trials where unrecognised unmeasured confounding is likely to be equally distributed between intervention and control groups, whereas unmeasured confounding will still be present using a cohort design. Furthermore, our estimate relates only to those women who are able to stop smoking as a result of the offer of financial rewards. There is a subset of women who are able to quit without this intervention. It is not known (and cannot be known) what their children’s birth weights would have been had they continued to smoke. These women are the most motivated (dubbed “independent quitters”)^12^ and their babies have greater birth weight than those of women who only quit due to the financial rewards. They likely adopted other lifestyle changes during pregnancy, such as dietary changes, which may confer additional benefits above those from stopping smoking.

The strength of this study is use of an important health outcome – birth weight - that has not been biased by the rewards process, as it is a routine measurement at birth in all jurisdictions.

The main weakness is the total sample size. In a randomised trial of the offer of financial rewards to stop smoking during pregnancy, only a minority of women will alter their behaviour as a result of the intervention. The majority will either continue to smoke, regardless of the rewards on offer, or would have stopped anyway, so the impact on birth weight at a population level will be heavily diluted. As an illustration, for a single trial to have 80% power at 5% significance to detect a mean 46g difference in birth weight between groups, assuming a standard deviation of 0.5kg, a total sample size of 3,712 participants would be required. Despite combining data from 8 trials, our maximum combined sample size was only 2,351.

### Is this result important?

Even though this result is statistically significant, is it important? ^26^ On the surface when judged by Cohen’s criteria it only indicates a small improvement measured in standard deviations of the outcome - birth weight (Cohen’s d approximately 0.1: 46g/500g). We therefore need to put the result into context: 1. In the current meta-analysis, smoking cessation was estimated to have a causal effect of increasing birth weight by 206g or 6.1% of average birth weight (3.4kg) for those compliant with the intervention who received rewards for stopping smoking, indicating a Cohen’s d of roughly 0.4 – medium importance (206g/500g); 2. Sceptics suggest that women ‘game’ the offer by stopping smoking for 24 hours to provide a normal carbon monoxide level to receive rewards ^27^. The improvement in birth weight shown in this review indicates that prolonged smoking cessation has taken place; 3. If Small for Gestational Age (SGA) births are reduced by 17.7% as suggested by this review then there will be one less baby born SGA for every 6 women who quit because of rewards; 4. The largest trial in the current meta-analysis was the most diverse with regards to pregnant smokers at least in the UK. The offer of financial rewards was ‘bolted-on’ to 7 very varied SSSs supporting pregnant women across three of the 4 UK countries ^28^. So, at least in the UK, offering financial rewards for smoking cessation during pregnancy is feasible. The outcome for this single multi-centre trial was a 50g increase in birth weight ^19^ comparable with the 46g improvement from the whole meta-analysis; 5. A meta-analysis of another already widely employed (in the UK) intervention for pregnant smokers, Nicotine Replacement Therapy, showed a comparable but non-significant increase in birth weight of 119g ^29^; 6. Offering financial rewards for smoking cessation during pregnancy is highly cost-effective providing £2 in health-care savings for every £1 extra spent on cessation support ^30^.

Further research, for example embedded in the funded countrywide roll-out of financial rewards for smoking cessation in pregnancy ^31^, is needed to clarify the most efficient frequency and level of financial rewards to employ.

## Conclusion

Financial rewards are being rolled out across England to help pregnant smokers to quit during pregnancy and to stay quit once their baby is born after recommendation from the Khan Review: Making Smoking Obsolete ^31^. Policy makers can be reassured that a biochemically measured increase in smoking cessation from the addition of financial rewards will be accompanied by an increase in birth weight and an overall reduction in health care costs.

## Contributor and guarantor information

DMT led and planned this study, wrote the PROSPERO protocol, applied for and gained ethics approval, contacted the authors of the previous systematic review and negotiated update of that review, made available data from one of the trials, attempted contact corresponding authors of trials included in the updated review to gather additional birth weight information and transferred this information for statistical analysis, accessed residual funding from one of the included trials to pay for JL to undertake the statistical analysis, made multiple drafts of this paper after comments from co-authors, submitted this paper for publication and accessed residual grant funding from FK to pay for open access publication. DMT is the corresponding author and guarantor.

JL undertook the statistical analysis for this paper, read and commented on multiple drafts of this paper.

AM supervised JL, helped write the PROSPERO protocol, developed the statistical analysis plan providing DMT with details of the additional birth weight information required, provided the additional information for one of the trials, read and commented on multiple drafts of this paper.

LK updated the previous systematic review, read and commented on multiple drafts of this paper.

STH allocated time for LK to update the systematic review through centre grant funding, made available additional information on 3 of the trials included in the updated review, gave advice on what additional birth weight data should be sought, read and commented on multiple drafts of this paper.

SHH supervised update of the previous review, allocated time for LK to update the systematic review through centre grant funding, agreed to make available data from one trial, read and commented on multiple drafts of this paper.

IB gave advice on additional birth weight data to access, made available data from one of the trials, read and commented on multiple drafts of this paper.

SJO made available data from one of the trials, read and commented on multiple drafts of this paper.

FK gave advice on the content of the discussion, commented on the manuscript multiple times and made available residual funding to pay for open access publication of this article.

IB worked closely with STH and SHH on 3 of the included trials, read and commented on multiple drafts of this paper.

LB supported DMT in all aspects of this study giving advice when required and providing overall supervision through long term liaison with FK, STH and IB.

DMT as corresponding author attests that all listed authors meet authorship criteria and that no others meeting the criteria have been omitted. As Corresponding Author DMT has the right to grant on behalf of all authors and does grant on behalf of all authors, a worldwide licence to the Publishers and its licensees in perpetuity, in all forms, formats and media (whether known now or created in the future), to i) publish, reproduce, distribute, display and store the Contribution, ii) translate the Contribution into other languages, create adaptations, reprints, include within collections and create summaries, extracts and/or, abstracts of the Contribution, iii) create any other derivative work(s) based on the Contribution, iv) to exploit all subsidiary rights in the Contribution, v) the inclusion of electronic links from the Contribution to third party material where-ever it may be located; and, vi) licence any third party to do any or all of the above.”

## Data Availability

Data is not available from this study

## Appendix 1: Table of excluded studies from Kock systematic review update searching from 17^th^ November 2022 to 5^th^ December 2023

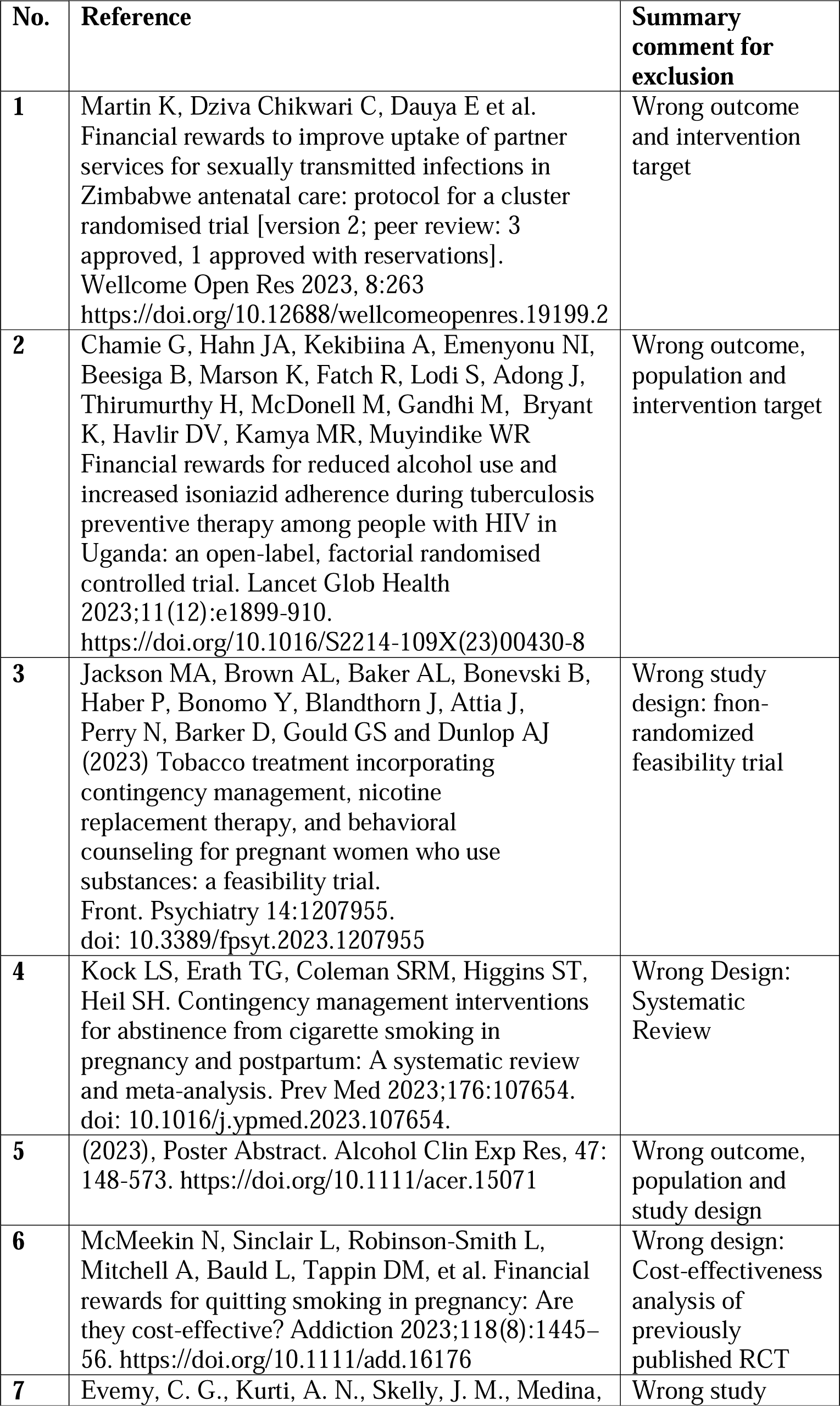
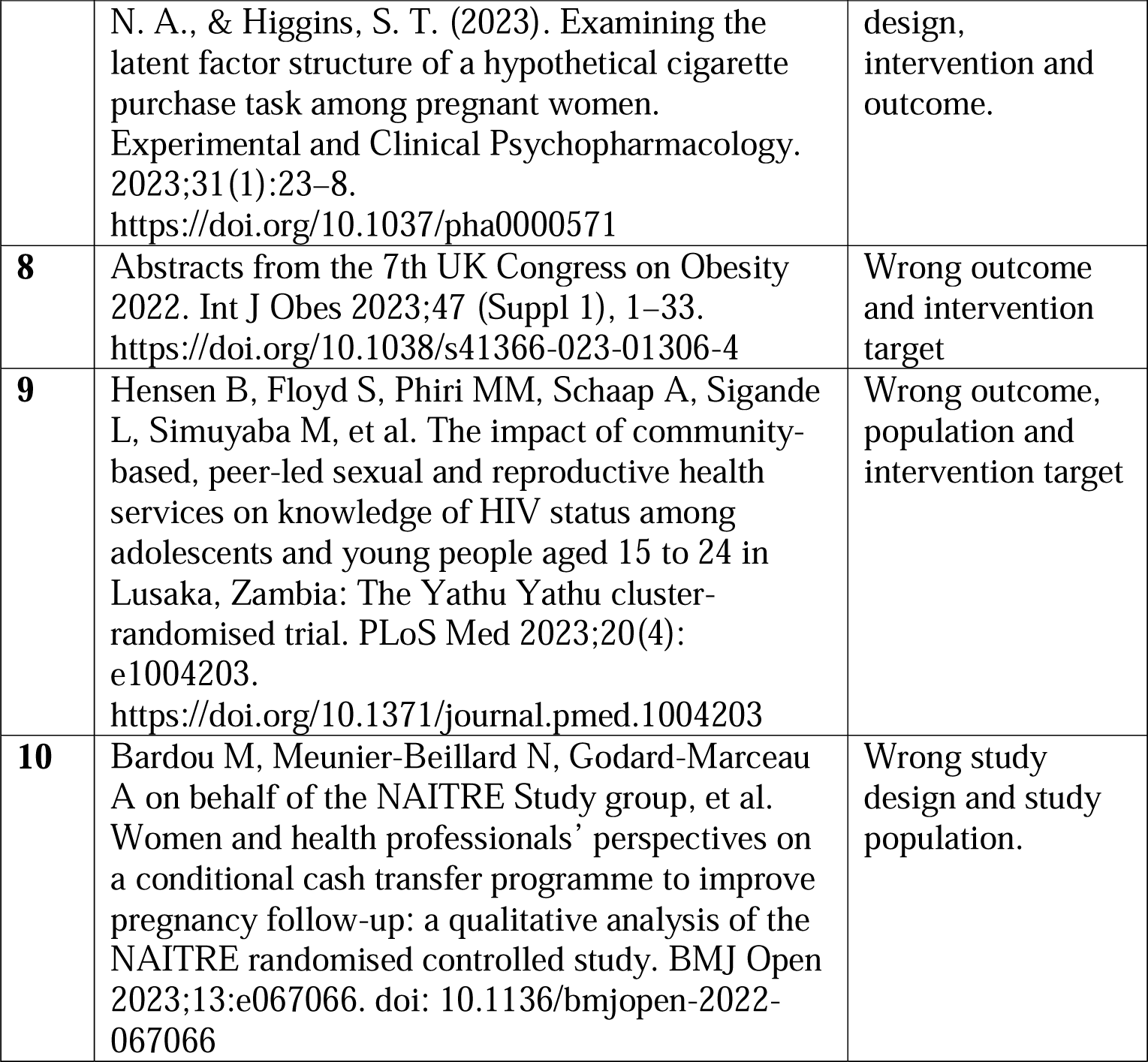

## Appendix 2: GRADE certainty of evidence table

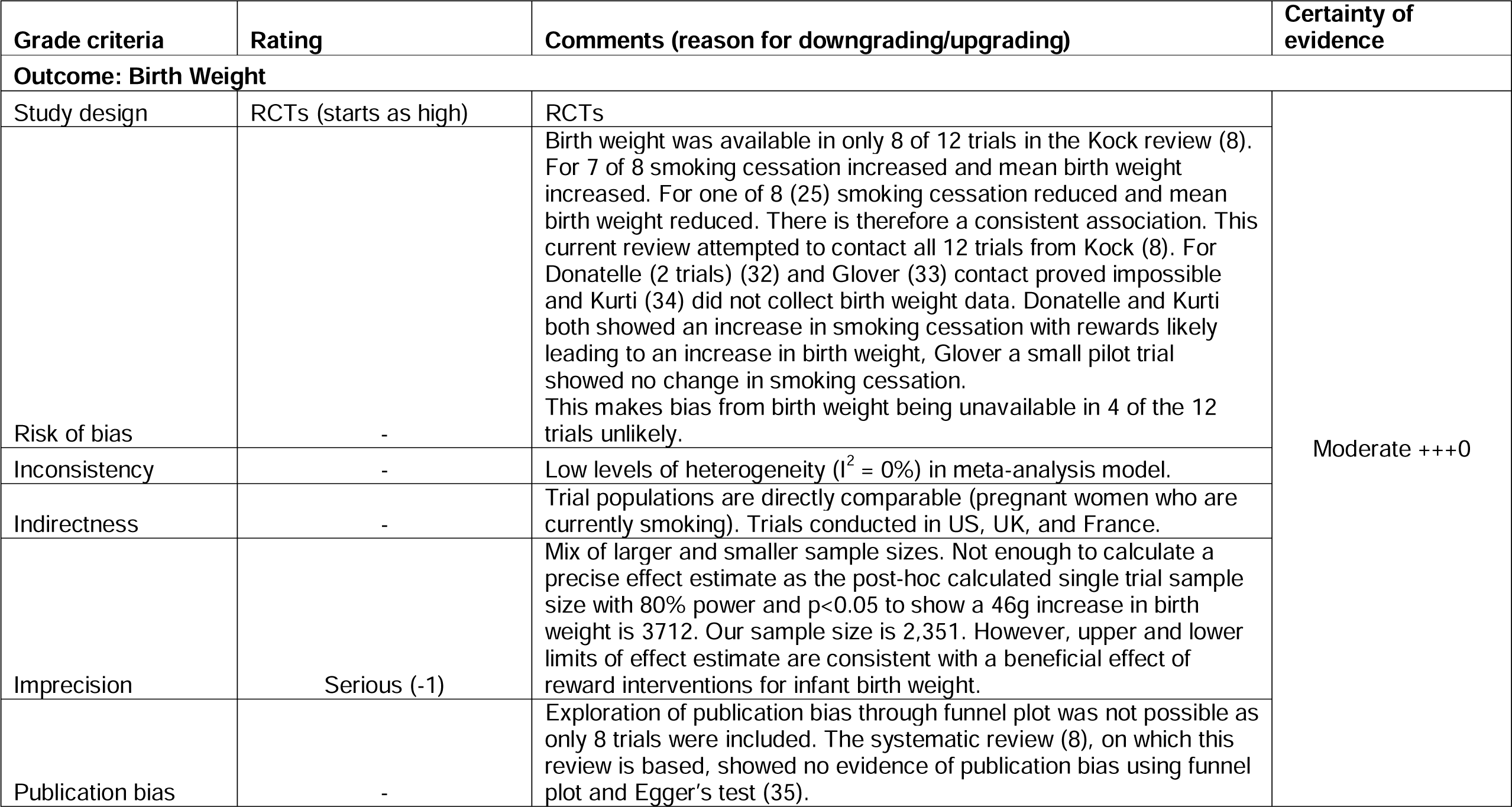

| Grade criteria | Rating | Comments (reason for downgrading/upgrading) | Certainty of evidence |
| --- | --- | --- | --- |
| **Outcome: Birth Weight** |  |  |  |
| Study design | RCTs (starts as high) | RCTs | Moderate +++0 |
| Risk of bias | - | Birth weight was available in only 8 of 12 trials in the Kock review (8). For 7 of 8 smoking cessation increased and mean birth weight increased. For one of 8 (25) smoking cessation reduced and mean birth weight reduced. There is therefore a consistent association. This current review attempted to contact all 12 trials from Kock (8). For Donatelle (2 trials) (32) and Glover (33) contact proved impossible and Kurti (34) did not collect birth weight data. Donatelle and Kurti both showed an increase in smoking cessation with rewards likely leading to an increase in birth weight, Glover a small pilot trial showed no change in smoking cessation. This makes bias from birth weight being unavailable in 4 of the 12 trials unlikely. |  |
| Inconsistency | - | Low levels of heterogeneity ($I^2 = 0\%$) in meta-analysis model. | |
| Indirectness | - | Trial populations are directly comparable (pregnant women who are currently smoking). Trials conducted in US, UK, and France. |  |
| Imprecision | Serious (-1) | Mix of larger and smaller sample sizes. Not enough to calculate a precise effect estimate as the post-hoc calculated single trial sample size with 80% power and $p < 0.05$ to show a 46g increase in birth weight is 3712. Our sample size is 2,351. However, upper and lower limits of effect estimate are consistent with a beneficial effect of reward interventions for infant birth weight. | |
| Publication bias | - | Exploration of publication bias through funnel plot was not possible as only 8 trials were included. The systematic review (8), on which this review is based, showed no evidence of publication bias using funnel plot and Egger's test (35). |  |

## What is known

Smoking during pregnancy is associated with miscarriage, stillbirth and babies born small for gestational age leading to lifelong poorer health outcomes.

Financial rewards for smoking cessation during pregnancy added to current Stop Smoking Service support more than doubles biochemically verified smoking cessation measured towards the end of pregnancy.

Offering financial rewards for smoking cessation during pregnancy is highly cost-effective providing £2 in health-care savings for every £1 extra spent on cessation support

## What this study adds

Policy makers can be reassured that spending limited resources on financial rewards for smoking cessation is associated with sustained cessation during pregnancy leading to an increase in infant birth weight.

There is some evidence that for every six women who give up smoking because they receive financial rewards for cessation during pregnancy one less baby will be born Small for Gestational Age.

