## Supplemental file for "Systematic review of randomised controlled clinical trials examining effectiveness of contingent financial rewards for smoking cessation during pregnancy: intention-to-treat and causal effects on birthweight"

**A systematic review update with data retrieval to allow meta-analysis and Complier Average Causal Effects (CACE) meta-analysis of birth weight.**

#### Supplementary material

David Tappin, Honorary Senior Research Fellow (https://orcid.org/0000-0001-8914-055X)^1^; Jiyoung Lee, Statistician^2^; Alex McConnachie, Professor of Clinical Trial Biostatistics^2^; Loren Kock, Statistician^3^; Stephen T. Higgins, Professor of Psychiatry, Professor of Psychological Science ^3^; Sarah H Heil, Professor of Psychiatry, Professor of Psychological Science ^3^; Ivan Berlin, associate professor of clinical pharmacology^4^; Steven J. Ondersma, Professor of Public Health^5^; Frank Kee, Professor of Public Health^6^; Ira Bernstein, Professor of Obstetrics, Gynecology and Reproductive Sciences^7^; Linda Bauld, Bruce and John Usher Professor of Public Health^8^.

^1^ Child Health, University of Glasgow, G12 8QQ, UK

^2^ Robertson Centre for Biostatistics, School of Health and Wellbeing, University of Glasgow, UK

^3^ University of Vermont, 1 South Prospect St, Burlington, VT 05401, United States.

^4^ Hôpital Pitié-Salpêtrière-Sorbonne Université, Paris, France

5 Department of Public Health, Michigan State University, 200 E. First St., Flint, MI, 48502.

^6^ Centre for Public Health, Queens University Belfast, Northern Ireland

^7^ Department of Obstetrics, Gynecology and Reproductive Sciences, Smith 410, UVMMC, 111 Colchester Avenue, University of Vermont Medical Center 05401

^8^ Usher Institute and SPECTRUM Consortium, University of Edinburgh.

#### Word count

Number of words: 664

Number of

Tables: 1

Figures: 11

References: 2

Correspondence to:

David Tappin, MD.

Child Health, University of Glasgow.

### Introduction

### Stop Smoking Services use biochemical verification of self-report of smoking cessation - such as carbon-monoxide breath test - as a measurable outcome, but some argue that participants receive financial rewards for smoking cessation during pregnancy by gaming the system – such as refraining from smoking for 24 hours prior to a carbon monoxide breath test (1). This study assesses birth weight, a measurement undertaken routinely by midwifery staff that will not have been affected by gaming.

### Methods

*From: PROSPERO 2024 CRD42024494262*

David Tappin, Loren Kock, Alex McConnachie, Linda Bauld, Jiyoung Lee. Financial incentives for smoking cessation during pregnancy: updating PROSPERO 2022 CRD42022372291 and re-focusing the outcome to birth weight. PRO Available from: https://www.crd.york.ac.uk/prospero/display_record.php?ID=CRD42024494262

*Search Strategy and selection criteria*

As outlined by Kock et al (2), ‘Medline, American Psychological Association PsycInfo, Embase, Cochrane (the Cochrane Central Register of Controlled Trials, the Cochrane Tobacco Addiction Group Specialised Register and the Cochrane Database of Systematic Reviews), and PubMed were searched from their inception until 5th December 2023 for published reports of RCTs or quasi-experimental or pragmatic trials (Cochrane, 2017) of incentives for abstinence from substance use among pregnant women. Only trials using an experimental design that will allow treatment effects to be attributed to the reward intervention were included.’ As described, Kock et al (2) originally planned to include RCTs of incentives for abstinence from substances other than tobacco, but due to a small number of studies (only two returned after screening), these were excluded and the review focused solely on interventions for tobacco smoking cessation.

The following search terms for published literature on contingency management interventions for abstinence from substance use during pregnancy were used.

Study design: RCT or randomi?ed controlled trial or trial or randomi?ed or controlled clinical trial or pragmatic clinical trial (title or abstract or keyword)

Contingency management interventions: contingency management or incentive or financial incentive or voucher (title or abstract or keyword)

Pregnant women: Pregnan* (title or abstract or keyword)

Substance use: cigarette or smoking or smok* or nicotine or opioid or opiate or drug or stimulant or cocaine or meth* or amphetamine or alcohol or dependence or substance or substance use*’

**Types of study to be included**

Randomised controlled trials (RCTs) allocating individuals to intervention or to control conditions.

**Types of interventions**

Incentive schemes to reward participants for validated cessation and abstinence in smoking cessation programmes.

Control groups could be usual care or a smoking cessation intervention similar to that provided in the experimental group, but without incentives. Studies comparing two interventions providing incentives, but which varied by the amount or type of incentive, or where intervention incentives were contingent on smoking cessation and control incentives were not contingent on smoking cessation, were also eligible.

**Condition or domain being studied**

Pregnancy and childbirth: Smoking cessation intervention during pregnancy

**Participants/population**

Pregnant women who are current smokers

**Intervention(s), exposure(s)**

The offer of financial incentives for smoking cessation during pregnancy in addition to routine stop smoking support

**Comparator(s)/control**

Routine stop smoking support

**Context**

Pregnant women who smoke tobacco usually in the context of maternity antenatal clinics but not excluding other contexts for the care of pregnant smokers

**Main outcome(s)**

Infant birth weight (in grams), gestation at birth (usually calculated from the date of the start of the Last Menstrual Period) and smoking cessation (biochemically verified) towards the end of pregnancy.

**Measures of effect**

Mean birth weight increase with the offer of financial incentives for smoking cessation in 4 groups for each included trial: 1. randomised to incentives for those who quit towards the end of pregnancy and 2. randomised to incentives who did not quit; 3. randomised to no incentives who quit towards the end of pregnancy and 4. randomised to no incentives who did not quit. Numbers in each group. Standard Deviation of birth weight for each group. Number of babies born in each group weighing less that 2500g.

**Additional outcome(s)**

We will request data from each trial on infants born small for gestational age (SGA) in the 4 groups above by asking each trial to provide data for mean and standard deviation Weight for Gestational Age z-score using the app: http://intergrowth21.ndog.ox.ac.uk/ and numbers in each group below the 10th percentile for gestational age [z-score < (-1.2816)].

**Measures of effect**

Effect measure will be Birth Weight for Gestational Age z score (using mean and Standard deviation z-score) and number below 10th percentile birth weight for gestational age [Small for Gestational Age: z-score < (-1.2816)] in each of the 4 groups.

**Supplementary table 1**

| Supplementary Table 1: Pooled estimates of the Intention-To-Treat (ITT) effects of the offer of financial rewards, and Complier Average Causal Effect (CACE) effects of smoking cessation, on study outcomes. Pooled estimates derived from random effects models. | | | |
| --- | --- | --- | --- |
| Outcome | Estimator | Analysis | Estimate (95% CI), p-value |
| Birth Weight (g) | Mean Difference | ITT | 46.3 (0.0, 92.6), p=0.0498 |
|  |  | CACE | 206.0 (-69.1, 481.1), p=0.1422 |
| Birth Weight <2.5kg | Risk Difference | ITT | -0.6% (-3.3%, 2.1%), p=0.6635 |
|  |  | CACE | -3.1% (-18.6%, 12.4%), p=0.6981 |
| Birth Weight z-score | Mean Difference | ITT | 0.016 (-0.122, 0.154), p=0.8163 |
|  |  | CACE | 0.275 (-0.380, 0.930), p=0.4110 |
| Small for Gestational Age | Risk Difference | ITT | -2.8% (-5.8%, 0.2%), p=0.0667 |
|  |  | CACE | -17.7% (-34.9%, -0.4%), p=0.0446 |

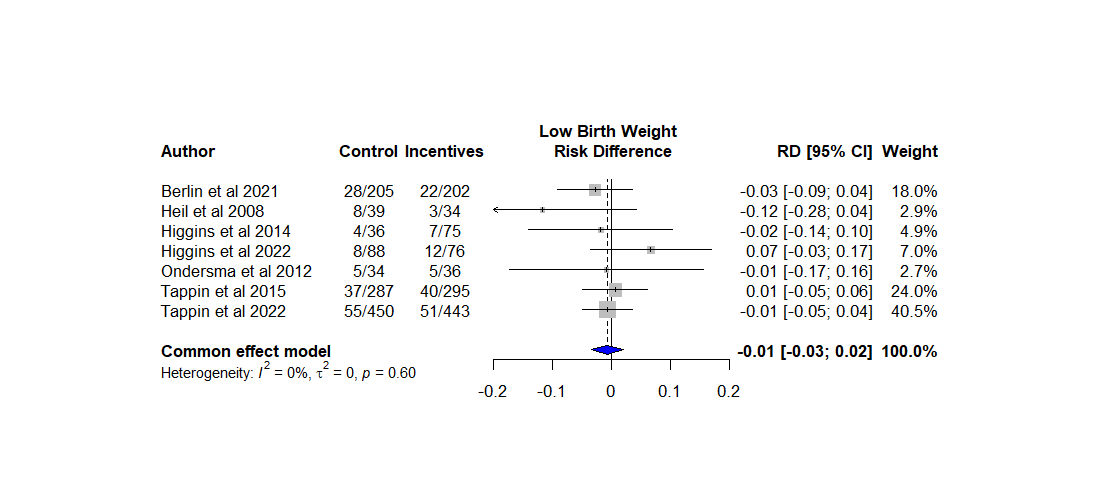

Supplementary Figure 1: Intention-to-treat (ITT) estimates of the effect of the offer of financial rewards for smoking cessation during pregnancy on low birth weight (<2.5kg), expressed as risk differences. The size of data markers is proportional to the weight in the meta-analysis.

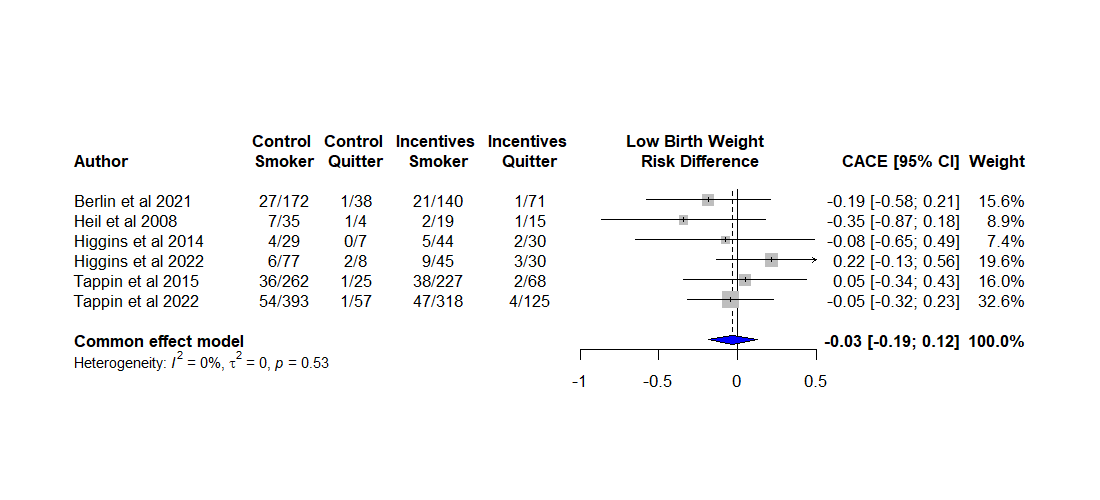

Supplementary Figure 2: Complier Average Causal Effect (CACE) estimates of the effect of smoking cessation during pregnancy on low birth weight (<2.5kg), expressed as risk differences. The size of data markers is proportional to the weight in the meta-analysis.

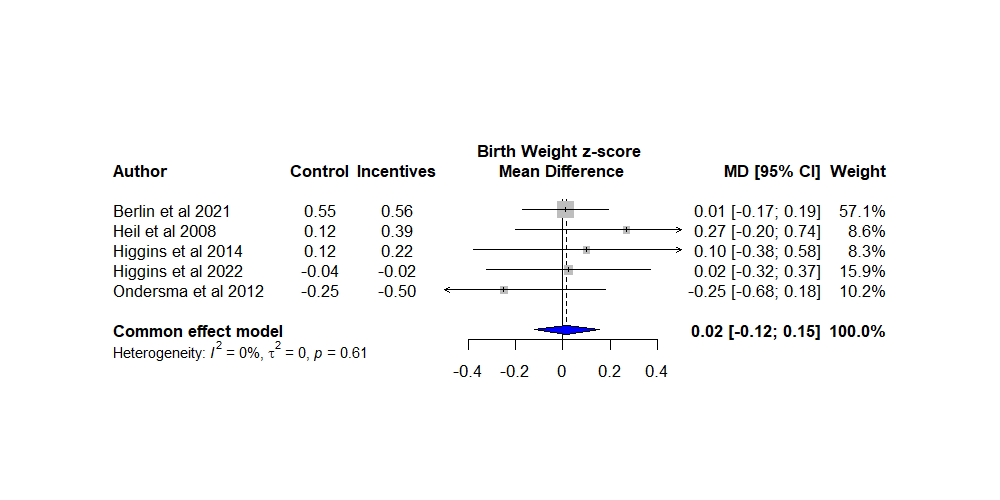

Supplementary Figure 3: Intention-to-treat (ITT) estimates of the effect of the offer of financial rewards for smoking cessation during pregnancy on birth weight z-score (adjusted for sex and gestational age). The size of data markers is proportional to the weight in the meta-analysis.

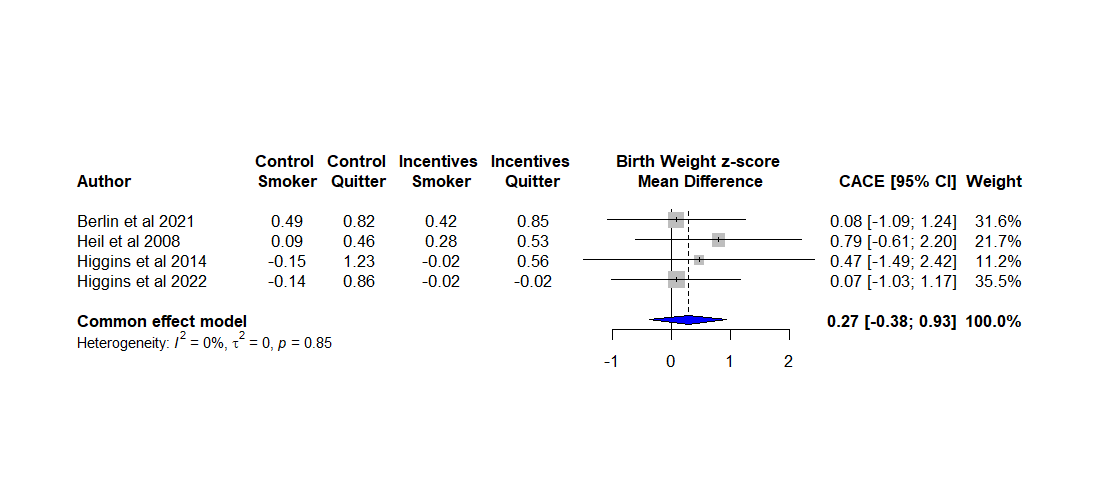

Supplementary Figure 4: Complier Average Causal Effect (CACE) estimates of the effect of smoking cessation during pregnancy on birth weight z-score (adjusted for sex and gestational age). The size of data markers is proportional to the weight in the meta-analysis.

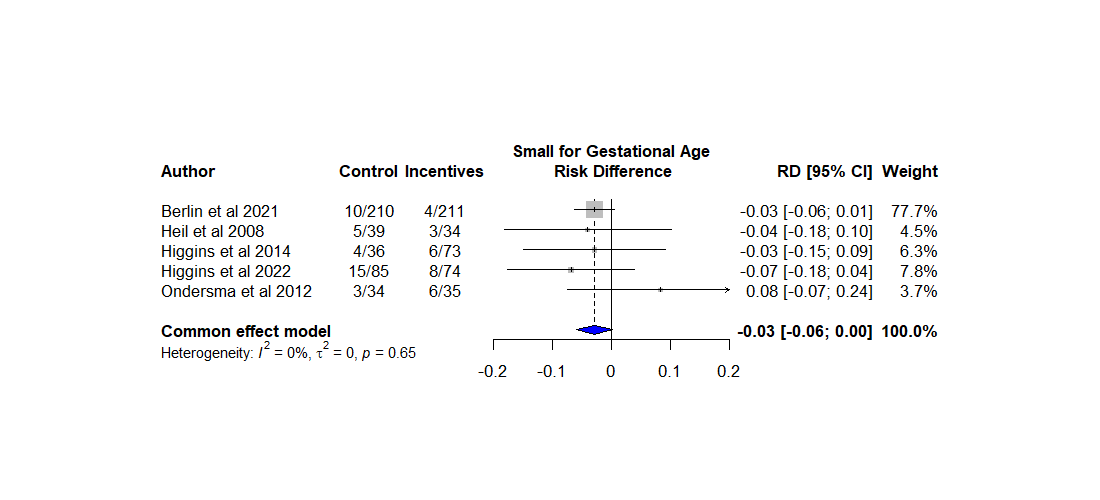

Supplementary Figure 5: Intention-to-treat (ITT) estimates of the effect of the offer of financial rewards for smoking cessation during pregnancy on risk of being born small for gestational age (<10^th^ percentile), expressed as risk differences. The size of data markers is proportional to the weight in the meta-analysis.

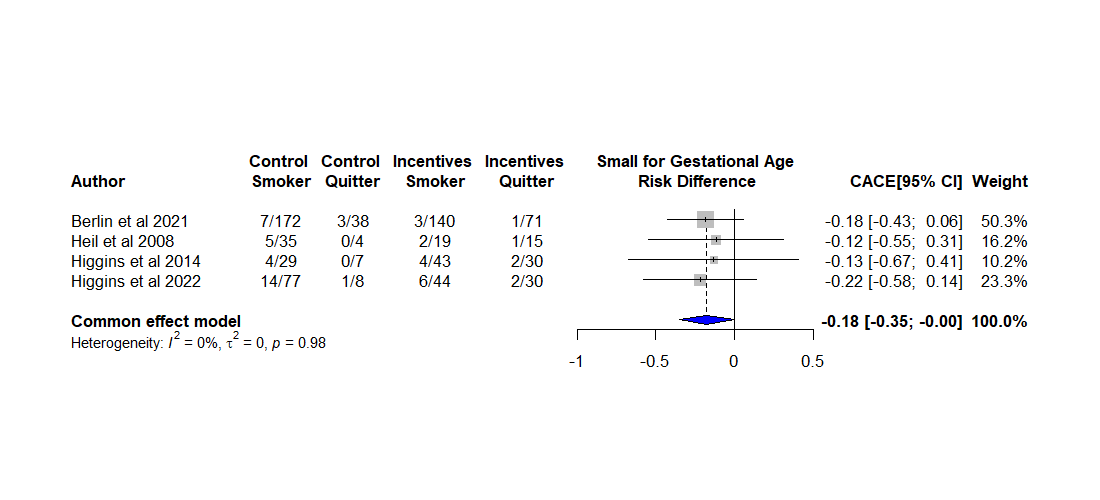

Supplementary Figure 6: Complier Average Causal Effect (CACE) estimates of the effect of smoking cessation during pregnancy on risk of being born small for gestational age (<10^th^ percentile), expressed as risk differences. The size of data markers is proportional to the weight in the meta-analysis.

| ITT | CACE |
| --- | --- |
| 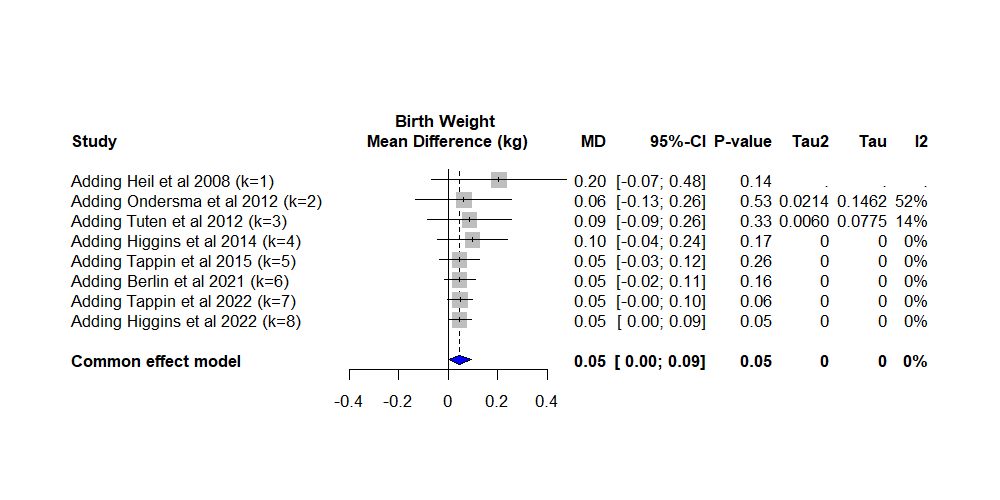 | 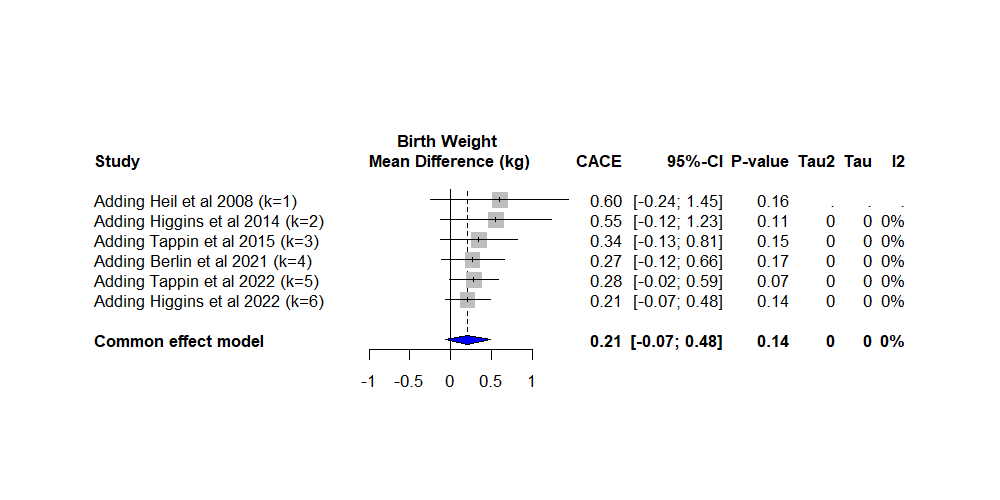 |
| 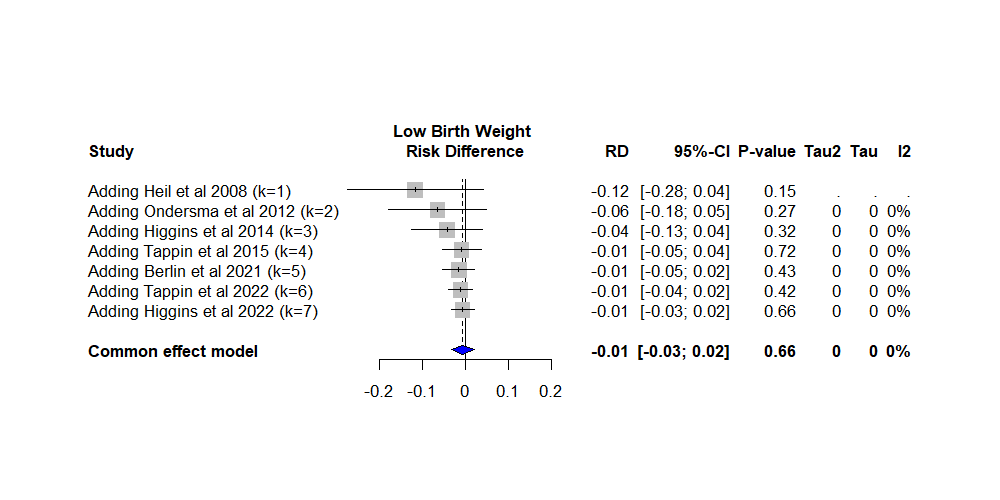 | 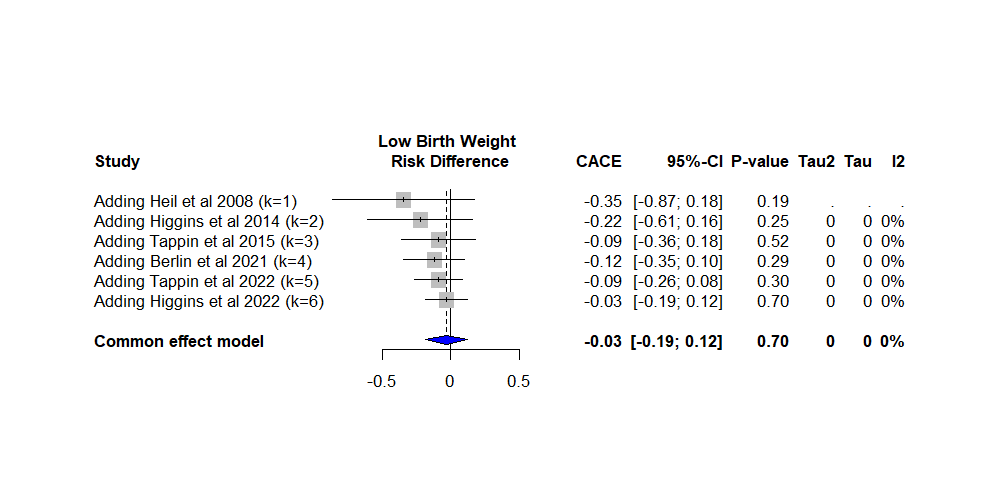 |
| Supplementary Figure 7: Cumulative meta-analyses of birth weight (top row), and low birth weight (<2500g, bottom row); left column shows ITT analyses (estimated difference between randomised groups), and right column shows CACE analyses (estimated causal effect of smoking cessation). Each plot shows the cumulative pooled estimate as results from each trial are added, in chronological order by publication date. | |

| ITT | CACE |
| --- | --- |
| 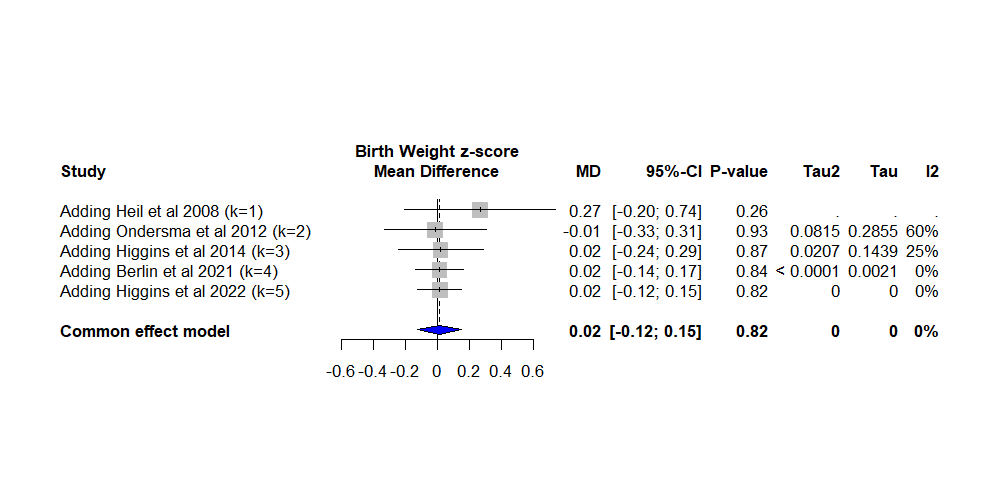 | 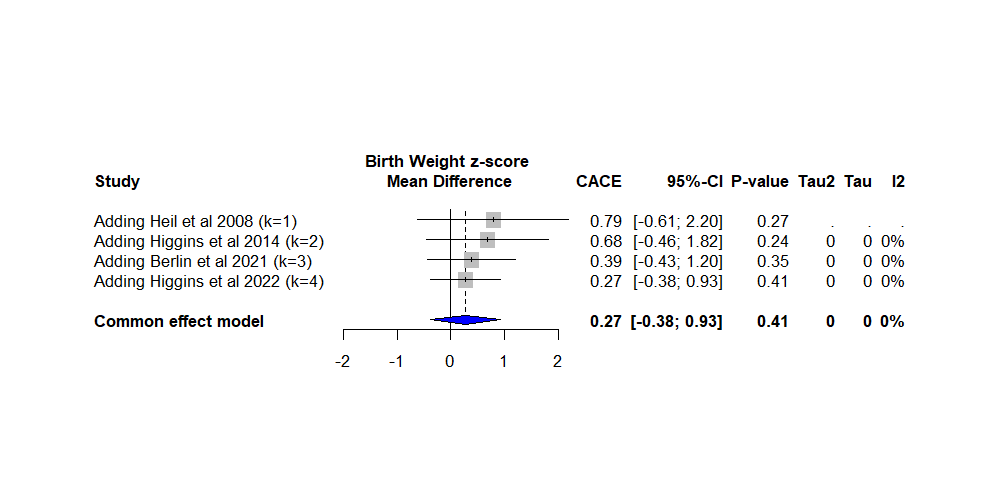 |
| 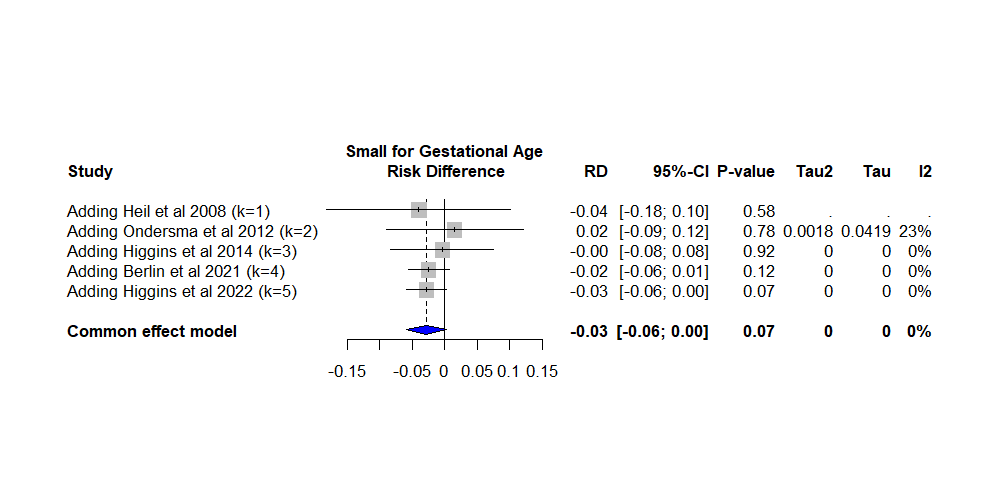 | 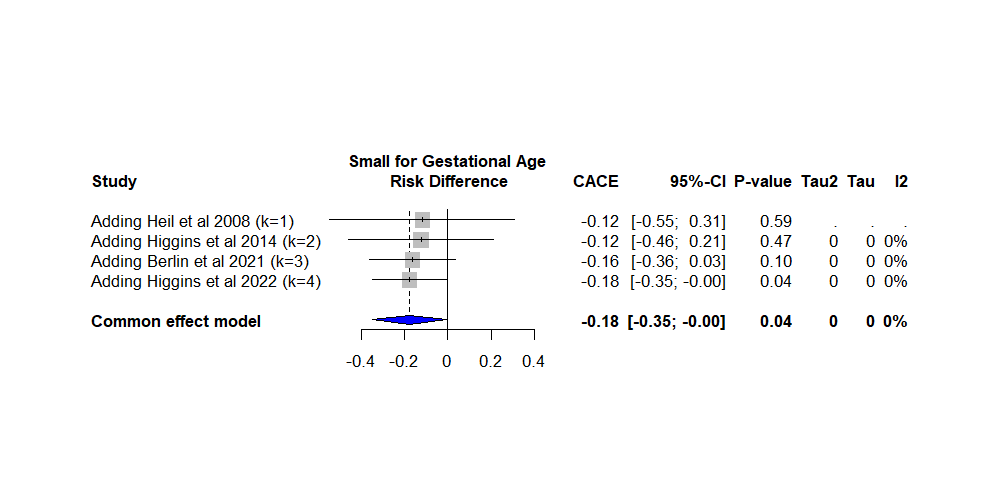 |
| Supplementary Figure 8: Cumulative meta-analyses of gestational age- and sex-adjusted birth weight z-score (top row), and small for gestational age (<10^th^ percentile, bottom row); left column shows ITT analyses (estimated difference between randomised groups), and right column shows CACE analyses (estimated causal effect of smoking cessation). Each plot shows the cumulative pooled estimate as results from each trial are added, in chronological order by publication date. | |

| ITT | CACE |
| --- | --- |
| 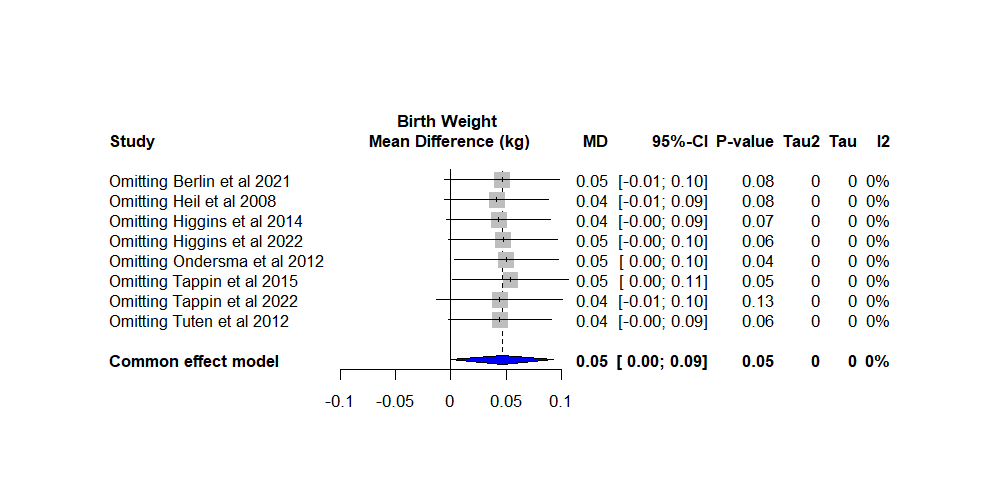 | 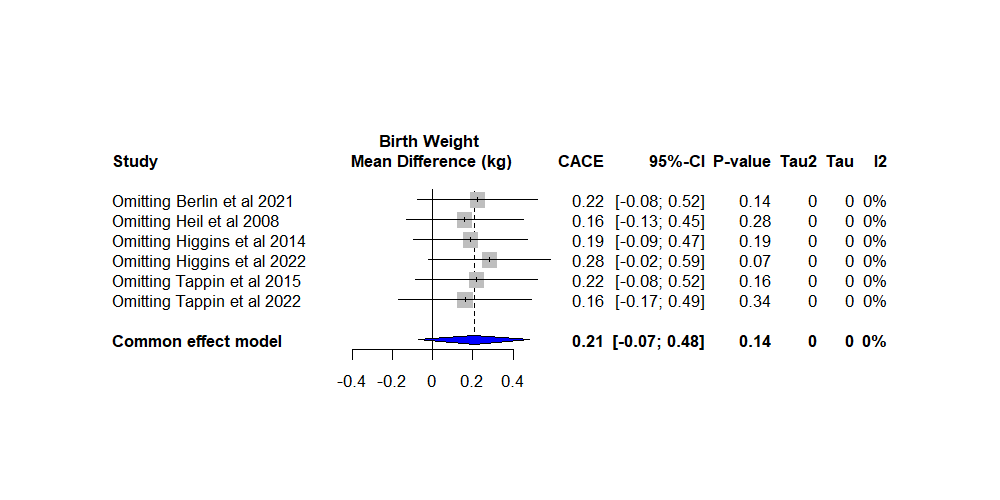 |
| 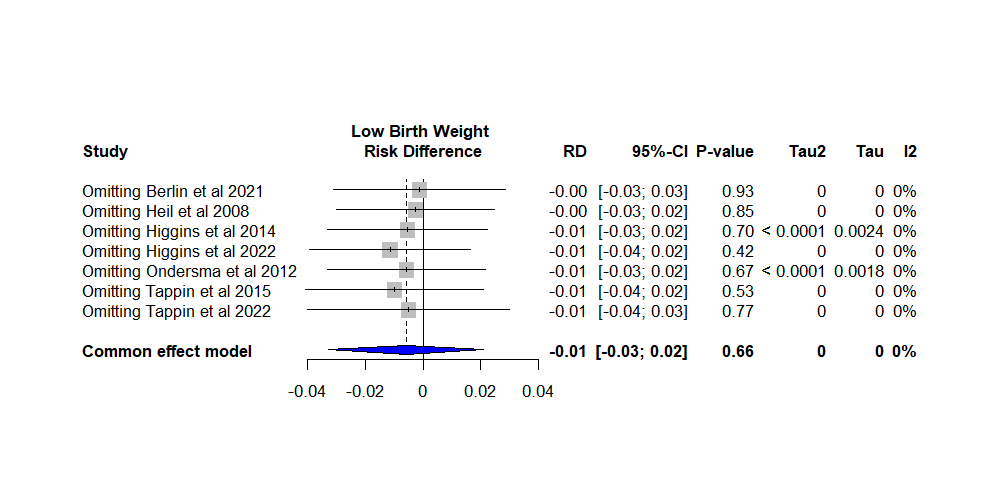 | 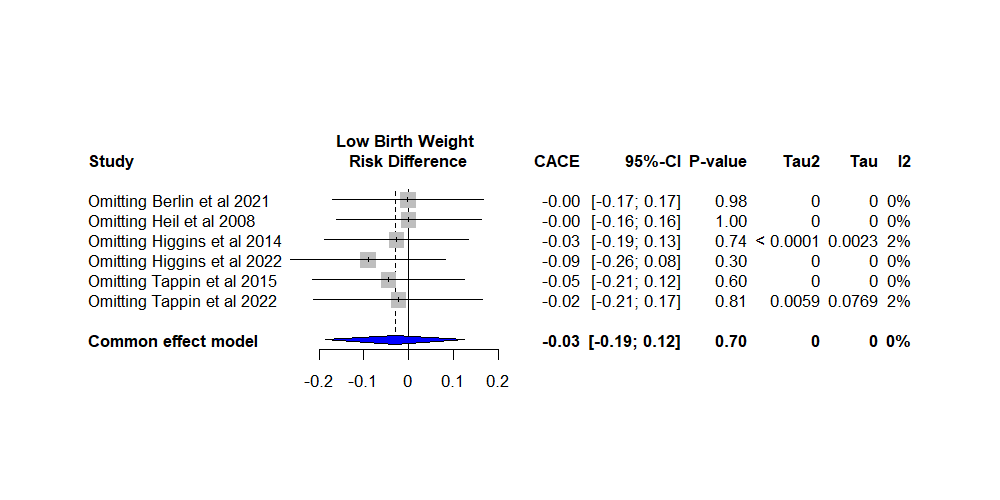 |
| Supplementary Figure 9: Leave-one-out meta-analyses of birth weight (top row), and low birth weight (<2500g, bottom row); left column shows ITT analyses (estimated difference between randomised groups), and right column shows CACE analyses (estimated causal effect of smoking cessation). Each plot shows the pooled estimate after the exclusion of each trial. | |
| ITT | CACE |
| 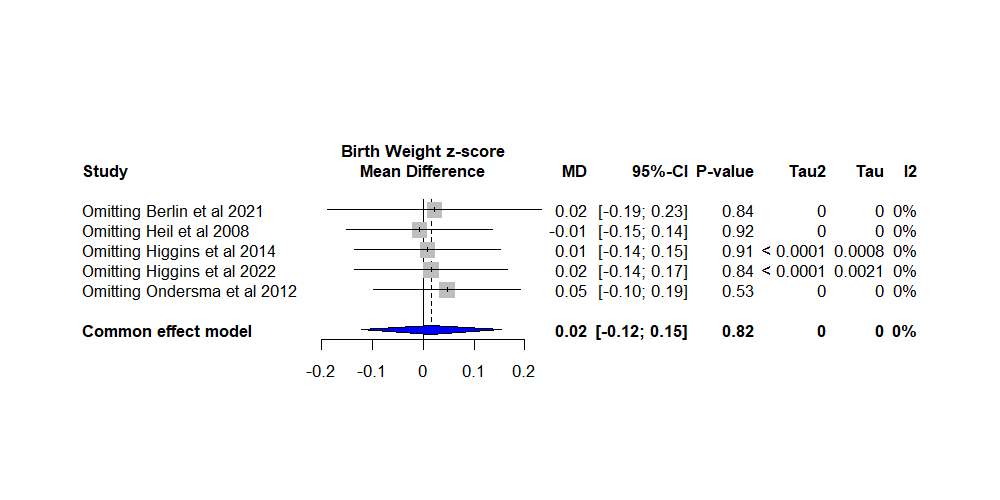 | 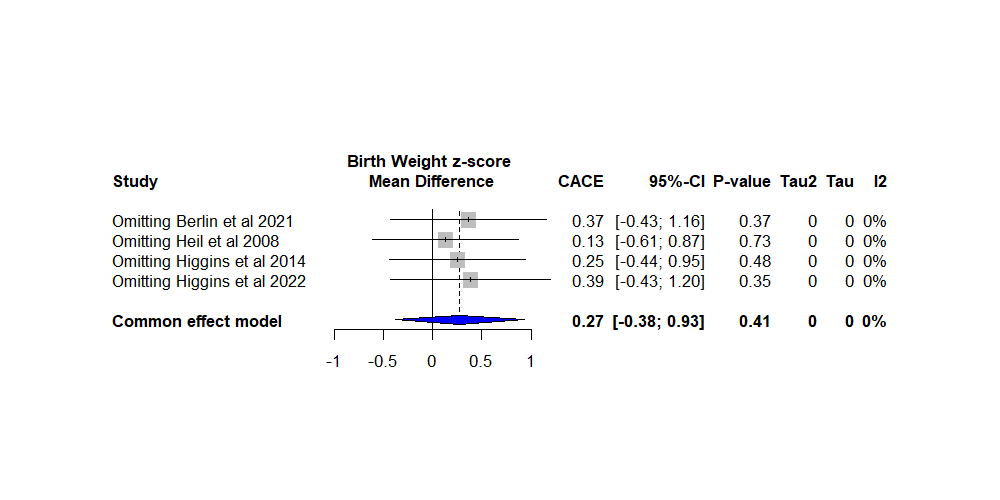 |
| 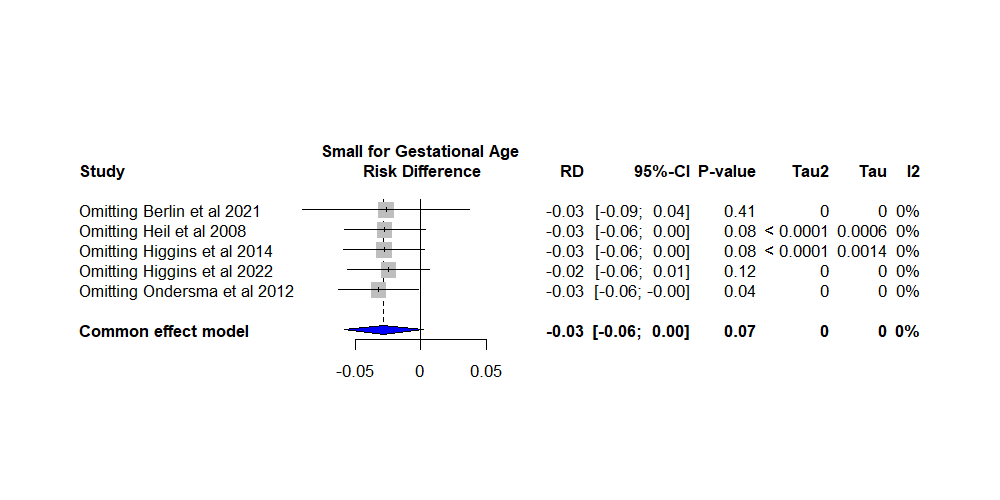 | 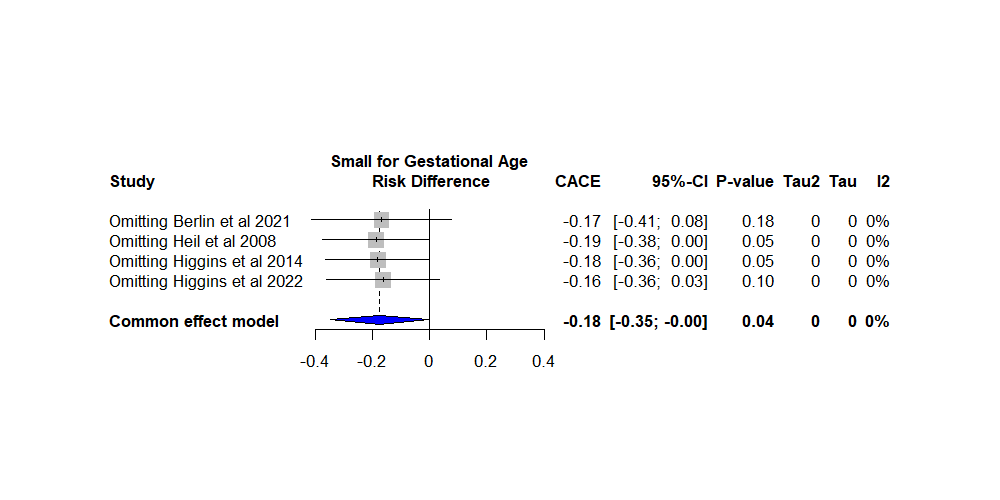 |
| Supplementary Figure 10: Leave-one-out meta-analyses of gestational age- and sex-adjusted birth weight z-score (top row), and small for gestational age (<10^th^ percentile, bottom row); left column shows ITT analyses (estimated difference between randomised groups), and right column shows CACE analyses (estimated causal effect of smoking cessation). Each plot shows the pooled estimate after the exclusion of each trial. | |

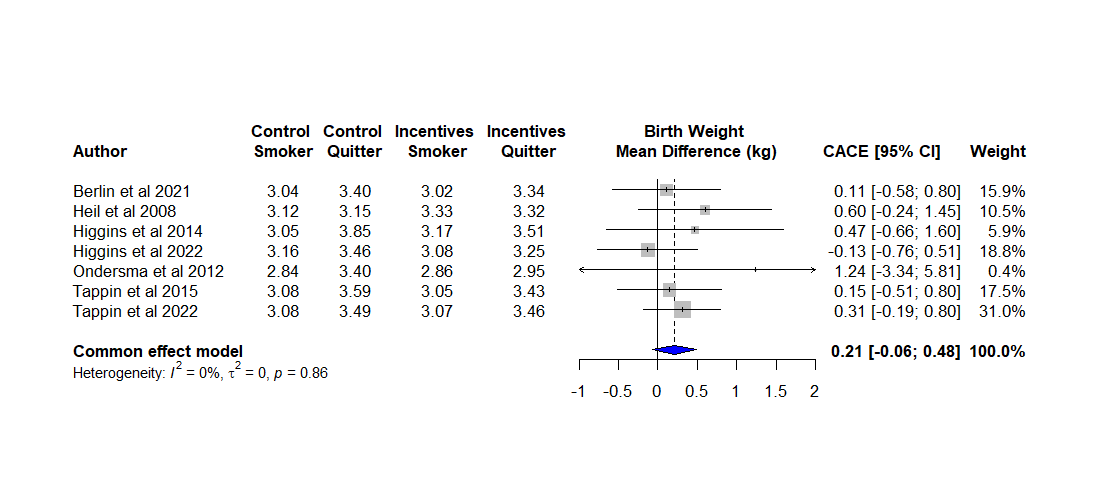

Supplementary Figure 11: Complier Average Causal Effect (CACE) estimates of the effect of smoking cessation during pregnancy on birth weight (kg). The size of data markers is proportional to the weight in the meta-analysis.
